# Diagnostic Machine Learning Applications on Clinical Populations using Functional Near Infrared Spectroscopy: A Review

**DOI:** 10.1101/2023.02.07.23285578

**Authors:** Aykut Eken, Farhad Nassehi, Osman Eroğul

## Abstract

Functional near-infrared spectroscopy (fNIRS) and its interaction with machine learning (ML) is a popular research topic for the diagnostic classification of clinical disorders due to the lack of robust and objective biomarkers. This review provides an overview of research on psychiatric diseases by using fNIRS and ML. Article search was carried out and 45 studies were evaluated by considering their sample sizes, used features, ML methodology, and reported accuracy. To our best knowledge, this is the first review that reports diagnostic ML applications using fNIRS. We found that there has been an increasing trend to perform ML applications on fNIRS-based biomarker research since 2010. The most studied populations are schizophrenia (n=12), attention deficit and hyperactivity disorder (n=7), and autism spectrum disorder (n=6) are the most studied populations. There is a significant negative correlation between sample size (>20) and accuracy values. Support vector machine (SVM) and deep learning (DL) approaches were the most popular classifier approaches (SVM = 20) (DL = 10). Eight of these studies recruited a number of participants more than 100 for classification. Change in oxy-hemoglobin (ΔHbO) based features were used more than change in deoxy-hemoglobin-based ones and the most popular ΔHbO-based features were mean ΔHbO (n=11) and ΔHbO-based functional connections (n=11). Using ML on fNIRS data might be a promising approach to reveal specific biomarkers for diagnostic classification.

## 1. Introduction

Subjective assessment criteria for psychiatric and neurological disorders are commonly used in clinics for diagnostic purposes. Questionnaires, self-reports, and clinical interviews are commonly used however, due to the subject-dependent nature of these measures that have always been considered a flaw in clinics (Pies, 2007). Diagnostic decisions are generally evaluated with objective measures such as laboratory tests or neuroimaging approaches. At this point, the usage of functional neuroimaging approaches as diagnostic tools is still widely being discussed (Henderson et al., 2020). Functional Magnetic Resonance Imaging (fMRI), Electroencephalography (EEG), Magnetoencephalography (MEG), Positron Emission Tomography (PET) and Functional Near Infrared Spectroscopy (fNIRS) are the most common functional neuroimaging approaches that are used to disclose potential biomarkers to discriminate psychiatric or neurological disorders having common symptoms or these disorders from healthy individuals (Nour et al., 2022).

As number of population-based neuroimaging datasets is getting increased over the years, due to its high-dimensional nature, researchers utilized machine learning (ML) methods for more advanced and individual-level analyses such as classification of disorders, prediction of clinical scores, or clustering of new subpopulations. ML applications in medicine gained great importance in recent years (Ahsan et al., 2022) and also in functional neuroimaging research (Bondi et al., 2023; de Filippis et al., 2019; Duffy et al., 2019; Rathore et al., 2017; Santana et al., 2022). Because, compared to conventional statistical approaches such as t-test, ANOVA, Kruskal-Wallis, or Friedman test, ML provides us with individual-level answers rather than average sense. This is quite remarkable in medicine. As we stated above (i) Many diseases/disorders/syndromes have common symptoms that make them complicated to distinguish each other by considering a single variable (ii) While diagnosing them, self-reporting of patients which is the conventional approach and also gold-standard for diagnosis of many disorders, might provide unreliable results due to having the potential to be easily manipulated. Therefore, there is a great necessity to reveal robust and objective biomarkers that provide individual accurate diagnosis (iii) In general, vast majority of behavioral and neuroimaging studies that focus on differences between patients and healthy individuals show these differences in average sense. However, these differences might not be valid for some individual cases due to huge variability across participants. At this point, the combination of neuroimaging approaches and ML techniques plays an important role in providing us some answers related to individual diagnoses rather than populations (Nenning & Langs, 2022). Previous reviews that cover a combination of ML techniques for the prediction of several diseases by using EEG (Craik et al., 2019), fMRI (de Filippis et al., 2019; Nakano et al., 2020) and PET (Duffy et al., 2019) showed that neuroimaging techniques and ML might have a future on individual diagnostic decisions.

Among these functional neuroimaging techniques, fNIRS is relatively new and promising approach due to its advantages (Baskak, 2018; Ehlis et al., 2014; Irani et al., 2007) and it has almost a contemporary history with artificial intelligence applications in medicine. However, due to lack of data and computational cost, ML usage in fNIRS studies was limited until recent years. After overcoming these limitations, ML usage has increased greatly through the last decade among fNIRS researchers. Compared to other neuroimaging modalities such as fMRI and PET, it is less expensive, portable, easy to apply and has more tolerance to motion artifacts. When compared to EEG, it has higher spatial resolution that allows the researchers to focus on a specific region of interest (ROI). In addition to these advantages, it also provides information about concentration changes of oxy-hemoglobin (ΔHbO), deoxy-hemoglobin (ΔHb) and total-hemoglobin (ΔHbT= ΔHbO + ΔHb) by using at least two different wavelengths. These advantages feature fNIRS as a potential alternative tool for the diagnosis of psychiatric diseases. It has widely been preferred by researchers and clinicians from many different fields such as infant development, cognition, anesthesia, motor control and psychiatric disorders (see review (Boas et al., 2014)).

Integrated fNIRS and ML systems should consist several systematic components as it is shown in Figure 1. A specific task or a resting-state procedure is conducted for data acquisition via a multi or single-channel fNIRS system. After data acquisition, a pre-processing step is carried out. In pre-processing step, several types of artifacts such as physiological noise (heartbeat, respiration, Mayer waves (Fekete et al., 2011a)), motion artifacts and very low-frequency noise (<0.1 Hz) need to be filtered out. For this purpose, band-pass filtering, signal detrending and motion artifact algorithms (Brigadoi et al., 2014) are used. Having carefully filtered the data, feature extraction is carried out. Feature extraction step directly affects the performance of classifiers. Due to this reason, a priori knowledge in either temporal or spatial behavior of hemodynamic response might be essential. Depending on the type of data (resting-state or task), extracted feature types might be different. Feature selection should also be carried out if the number of features is high. This may lead to a dimensionality problem which may cause an overfitting or underfitting problem. In this step, there are several algorithms that might be used such as Principal Component Analysis (PCA), Least Absolute Shrinkage and Selection Operator (LASSO), t-test and Recursive Feature Elimination (RFE). Cross-validation types (Hold-Out, Leave-one-out (LOOCV) and K-fold) are generally selected depending on the amount of data and expected computational cost. In some studies, hyperparameter optimization techniques such as grid-search, random-search or Bayesian are used to improve the performance of classifiers or predictors. For classification or prediction, methods such as Support Vector Machine (SVM), K-nearest neighborhood (KNN), linear discriminant analysis (LDA), Gaussian process classifier (GPC), Random Forest (RaF), Linear regression (LR) and Convolutional Neural Network (CNN) as a deep learning model are used.

**Figure 1.**
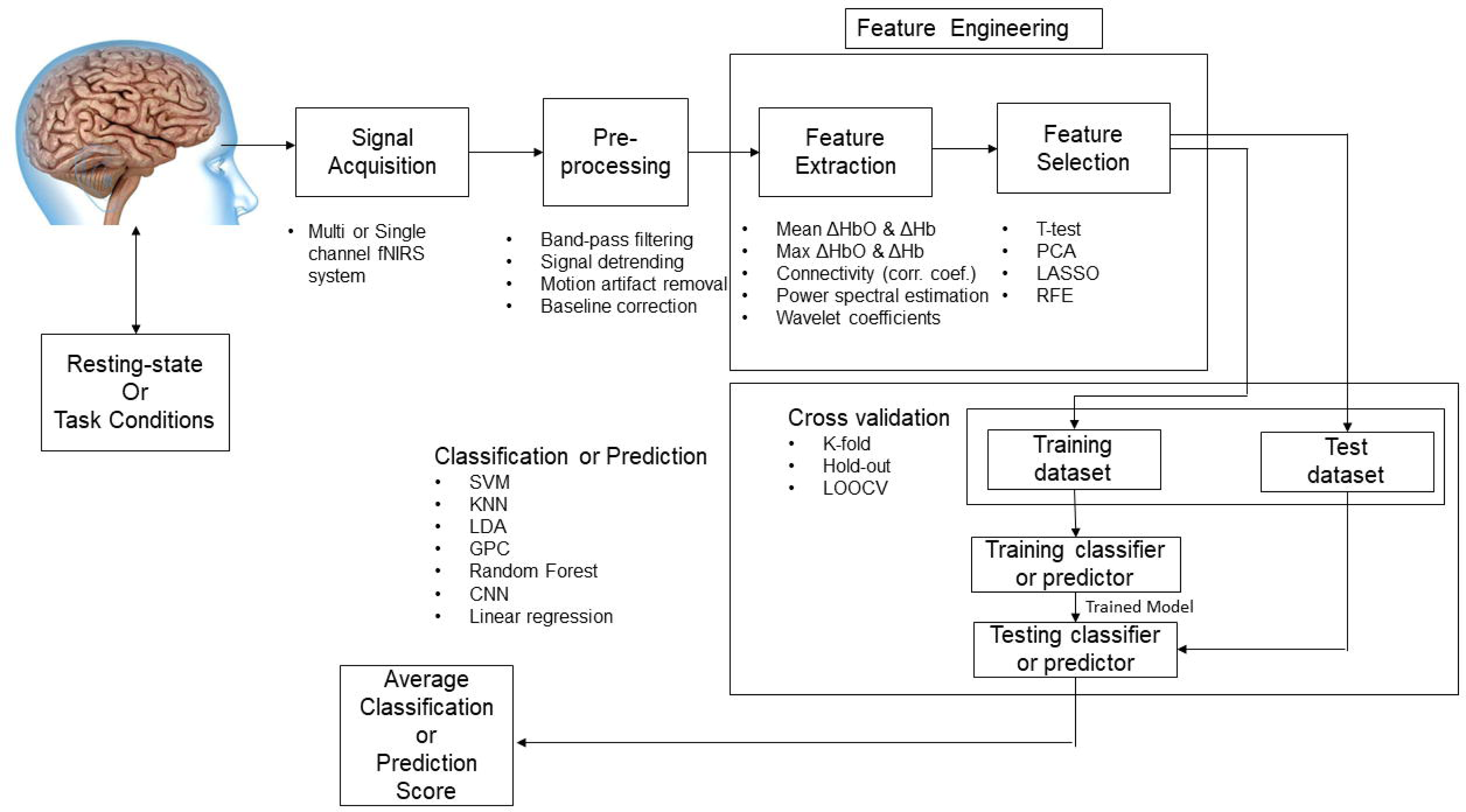
A general pipeline for classification or prediction of a clinical disease or disorder. fNIRS: Functional Near Infrared Spectroscopy, ΔHbO : Oxy-hemoglobin concentration change, ΔHb: Deoxy-hemoglobin concentration change, PCA : Principcal Component Analysis, LASSO: Least Absolute Shrinkage and Selection Operator, RFE : Recursive Feature Elimination, LOOCV: Leave-one-out cross validation, SVM: Support Vector Machine, KNN: K-nearest neighborhood, LDA: Linear Discriminant Analysis, GPC: Gaussian process classifier, CNN: Convolutional Neural Network.

Our primary objective to review fNIRS-based ML studies is to provide a general overview the potential of fNIRS and ML to assess psychiatric disorders and provide an insight to researchers about to the classification strategies, potential features to related disorders. We also discussed potential problems usage of fNIRS for diagnostic purpose and suggest questions for further studies. This review includes a general overview of these applications on clinical populations. To our best knowledge, this is the first review that covers machine learning studies diagnosing psychiatric disorders using fNIRS. There is a recent review focusing on deep learning applications using fNIRS data including cortical analysis, preprocessing, BCI and diagnostic applications (Eastmond et al., 2022). However, as we stated above we also discussed the features that can be considered as potential biomarkers.

## 2. Materials and Methods

### 2.1. Identification

The present study was performed according to the “Preferred Reporting Items for Systematic reviews and Meta-Analyses” (PRISMA) statement (Page et al., 2021), shown as a schema in Figure 2. The search procedure was initiated by using Web of Science and PubMed databases. We used the keywords (“Functional Near Infrared Spectroscopy” OR “Near Infrared Spectroscopy” OR “Diffuse Optical Imaging”) AND (“Machine Learning” OR “Prediction” OR “Classification”) that describe in Table 1 in detail. Original research papers published from starting 2010 until end of December 2022 were included. A total of 1552 (Pubmed: 852, Web of Science:705) search results that were published in Science Citation Indexing and Science Citation Indexing-Expanded, were reached. After removing the duplicate results, 1500 articles were left. Articles Conference proceedings and reviews excluding, 1459 articles were left. We also excluded the clinical state based studies (classification of pain, stress, anxiety conditions), non-clinical studies, brain-computer interface (BCI) studies and studies closely related to BCI such as motor and mental arithmetic tasks since it has been extensively reviewed by Naseer and Hong (Naseer & Hong, 2015). Among these studies, we also excluded the studies that either the accuracy value was not clearly reported or had accuracy values lower than %60.

**Figure 2.**
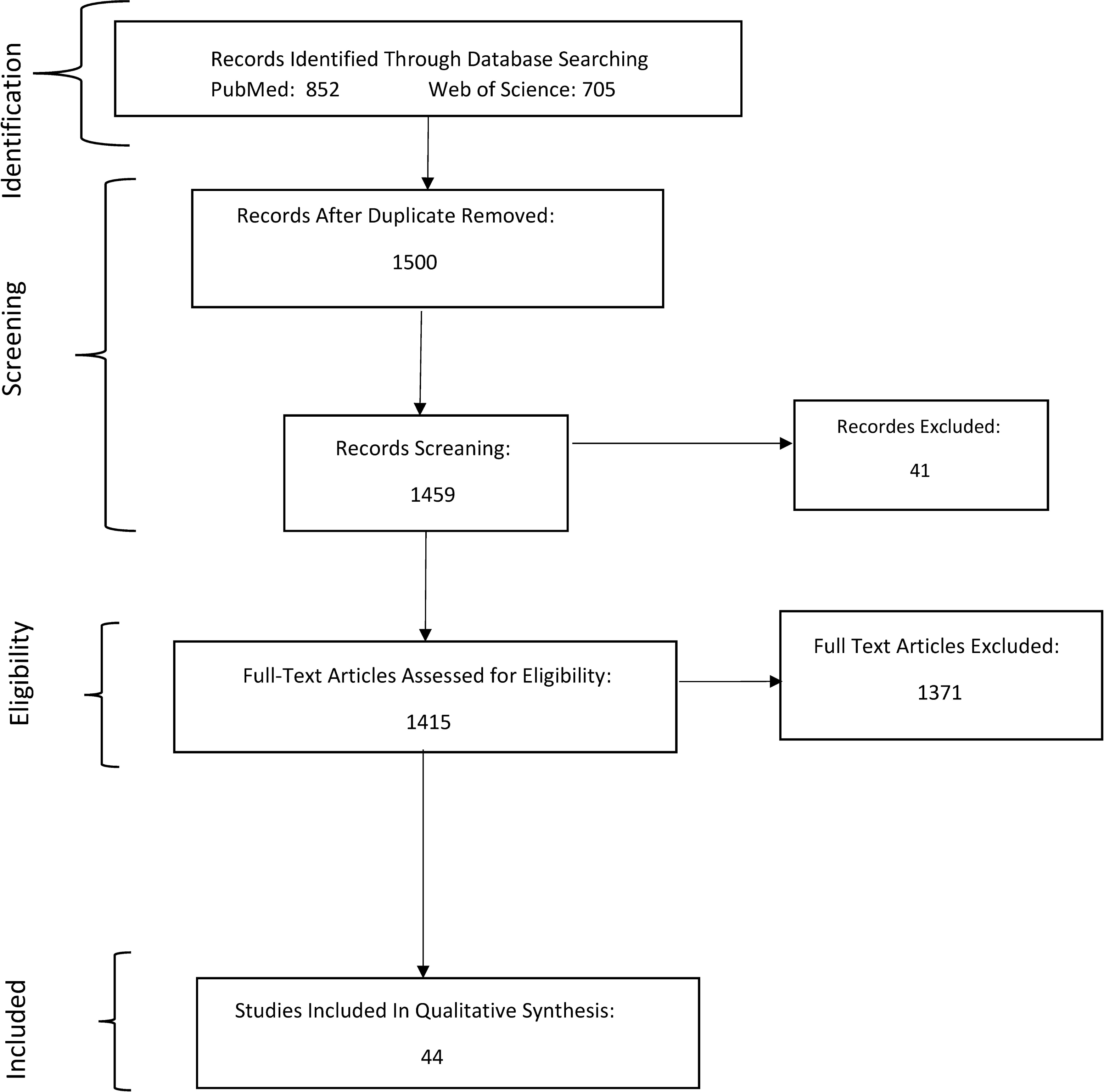
PRISMA flow chart that was followed in this review.

**Table 1:** Utilized databases and search terms.

| Database Name | Searching words |
| --- | --- |
| **Pubmed** | (classification[Title/Abstract] OR machine learning[Title/Abstract] OR prediction[Title/Abstract]) AND (functional near infrared spectroscopy[Title/Abstract] OR near infrared spectroscopy[Title/Abstract] OR diffuse optical imaging[Title/Abstract]) |
| **Web of Science** | (TI=(classification OR machine learning OR prediction)) AND TI=(functional near-infrared spectroscopy OR near-infrared spectroscopy OR diffuse optical imaging) |

### 2.2. Screening and Inclusion

We scanned and reported 45 articles that were suitable for our context. All included studies are summarized in Table 2. Extracted data types from publications were first author and year of the publication, populations, objective of the study, experiment type (task/resting), used fNIRS system, region of interest with 10-20 position if available, sample size, used features to train and test the model, used machine learning algorithm, cross-validation technique, hyperparameter optimization type, obtained the highest accuracy, other classification scores and comments related to the study. Studies were grouped according to the focused clinical population. For some studies, two different populations were studied such as Schizophrenia (SCZ), Bipolar Disorder (BP) vs Healthy Controls (HC) (Eken et al., 2022), Alzheimer’s Disease (AD), Mild Cognitive Impairment (MCI) and HC (E. Kim et al., 2021; J. Kim et al., 2022) and two different group of SCZ (Azechi et al., 2010). These studies were included twice for each clinical population and in total 49 studies were considered. In addition this, we added a narrative review of included studies for every disorder separately and added graphical information to discuss critical points in the literature.

**Table 2.**
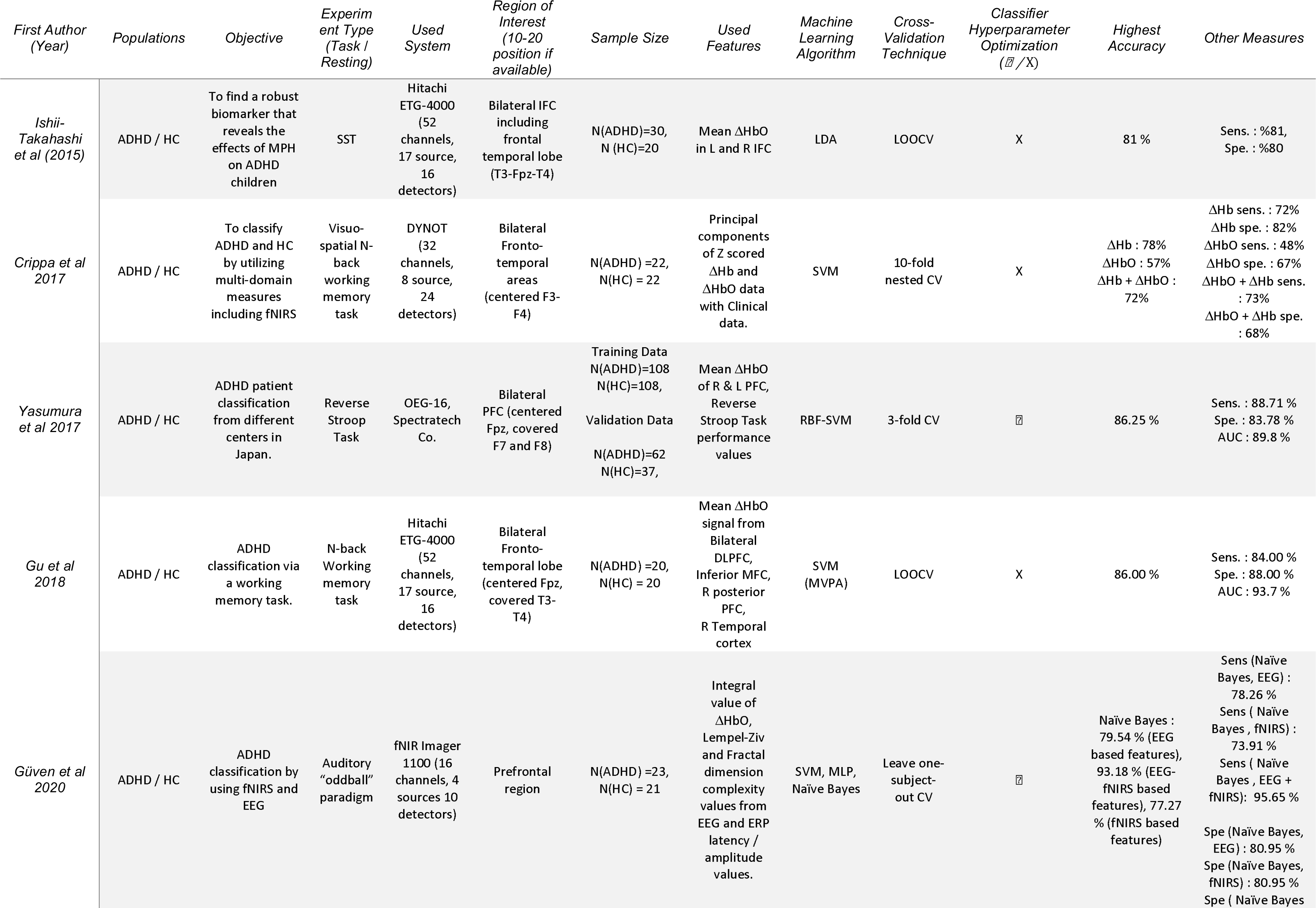
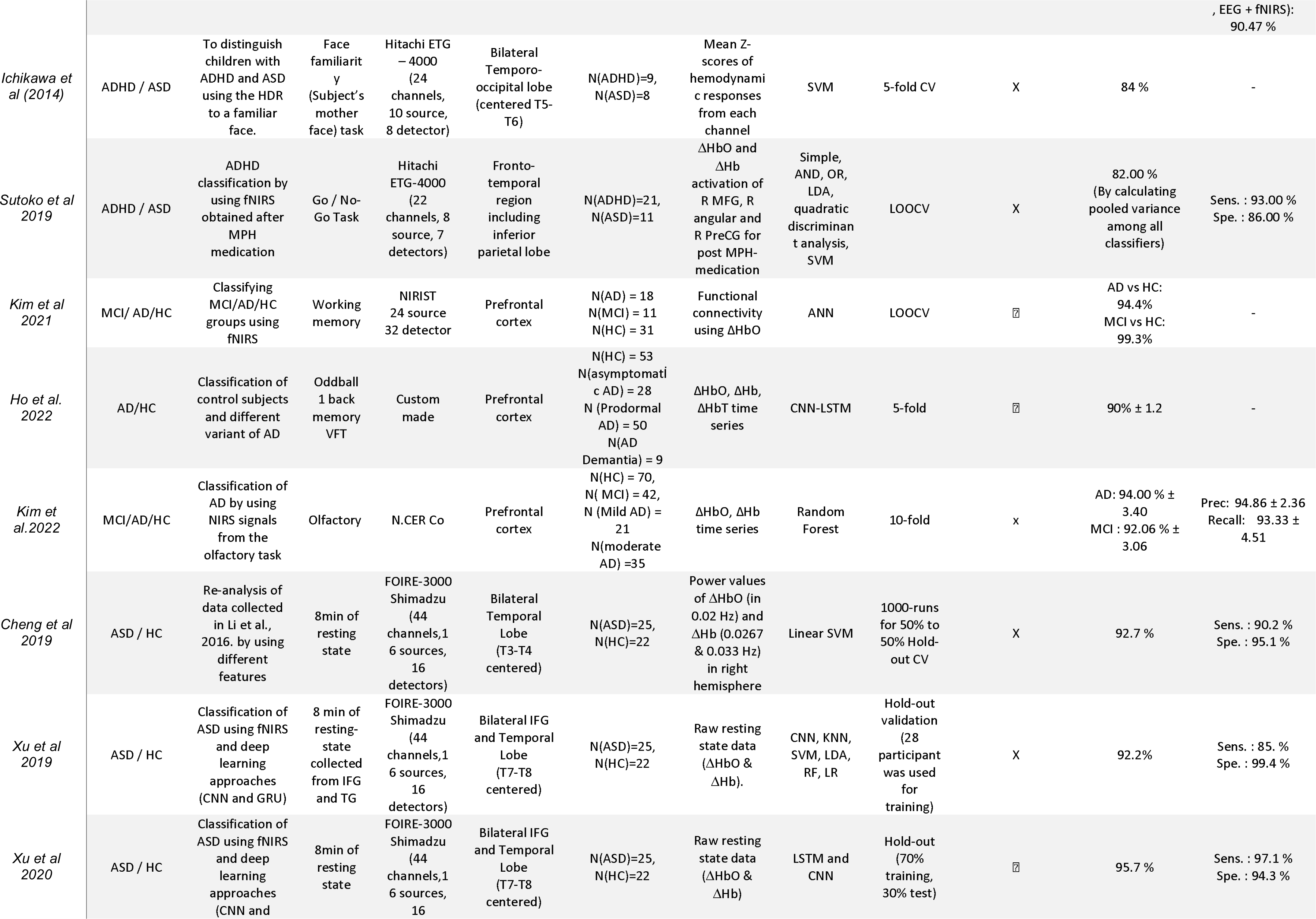
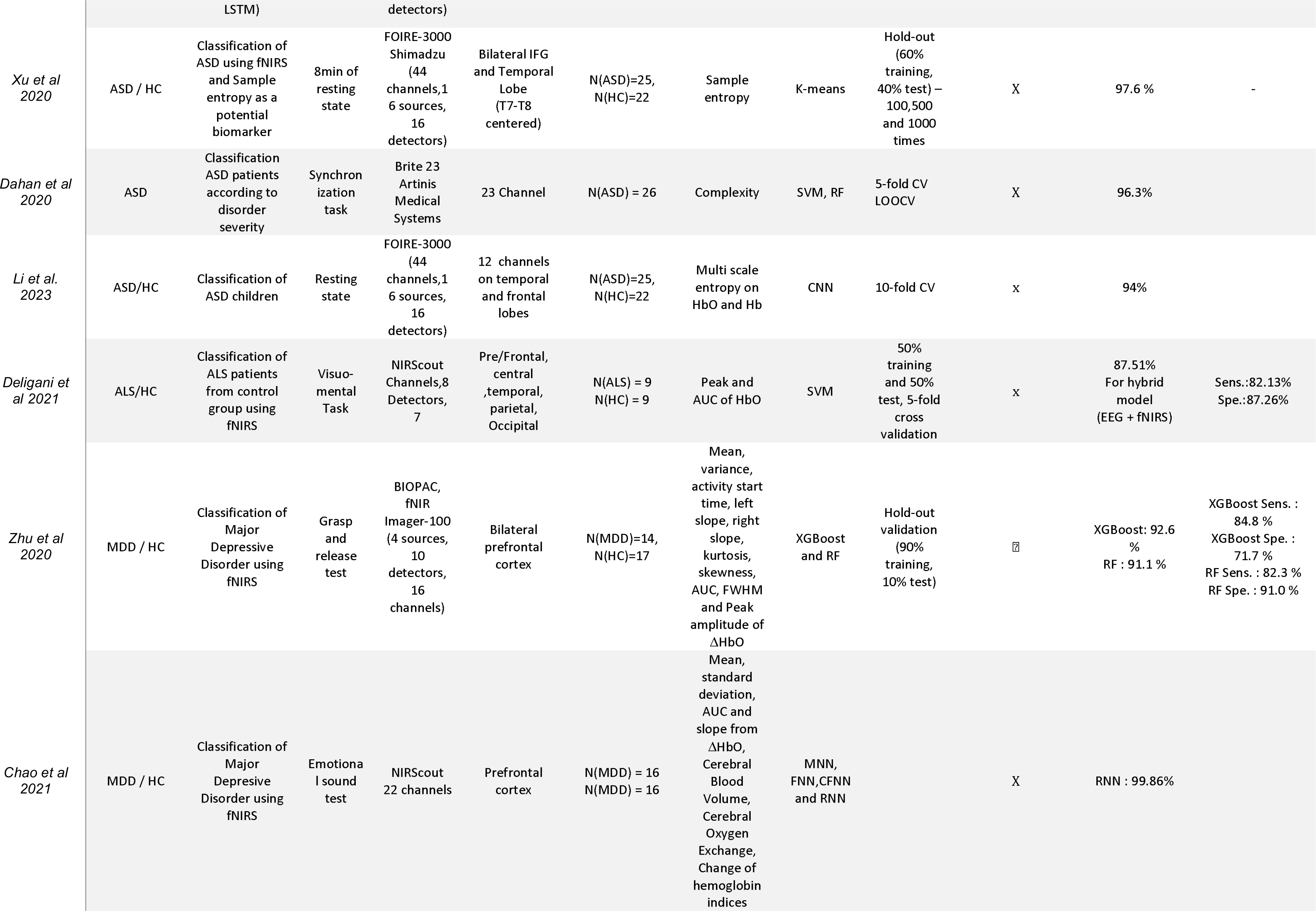
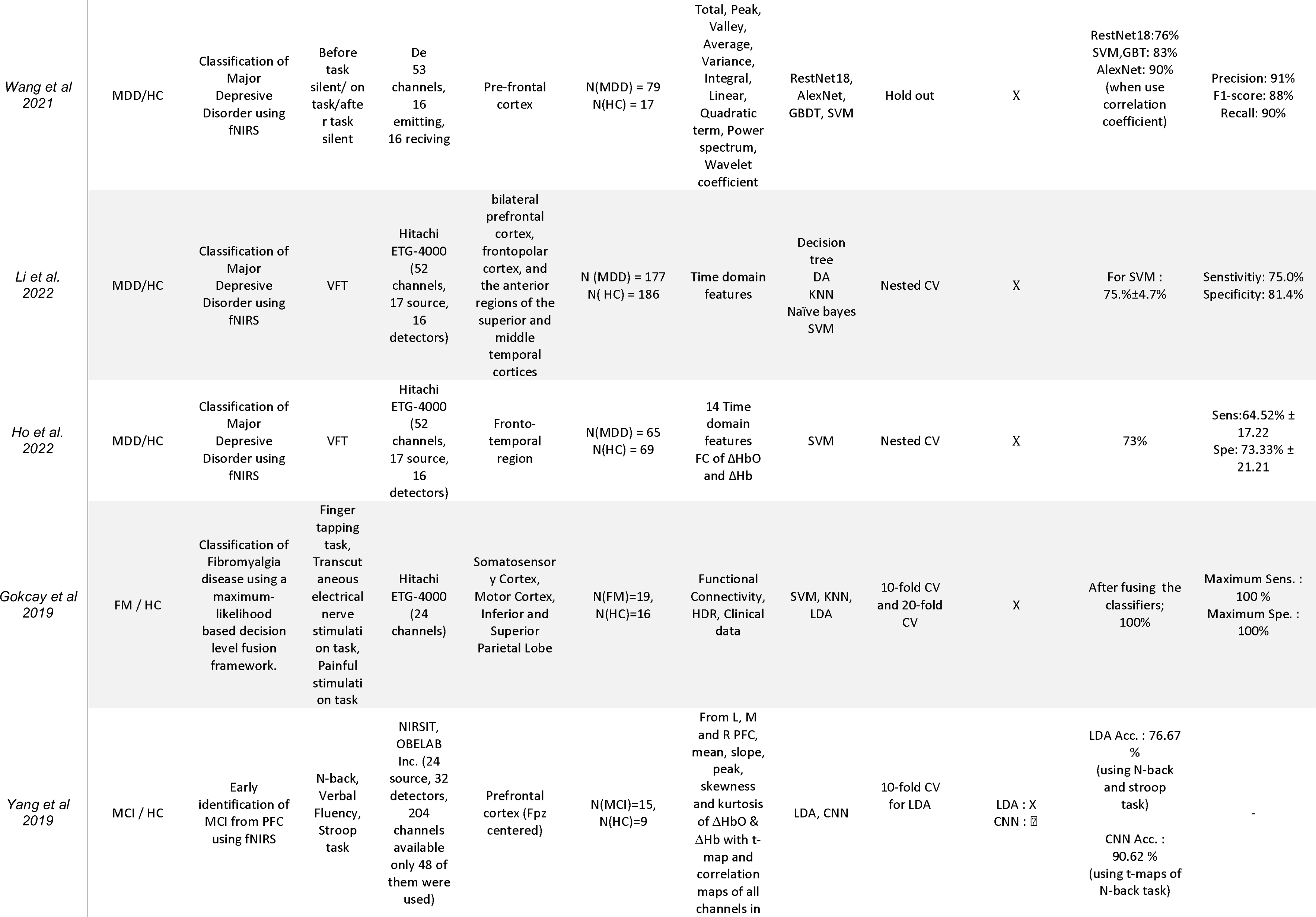
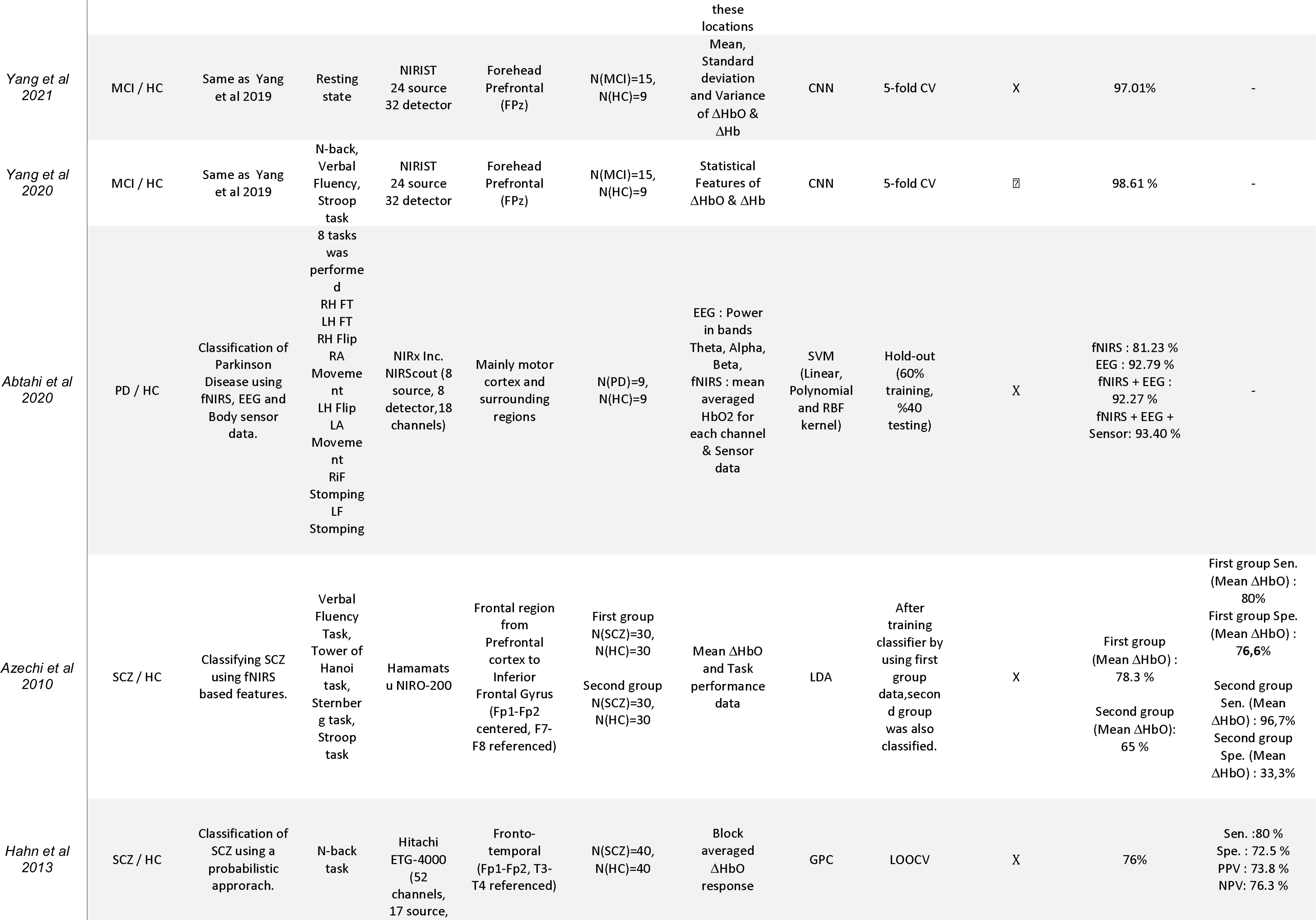
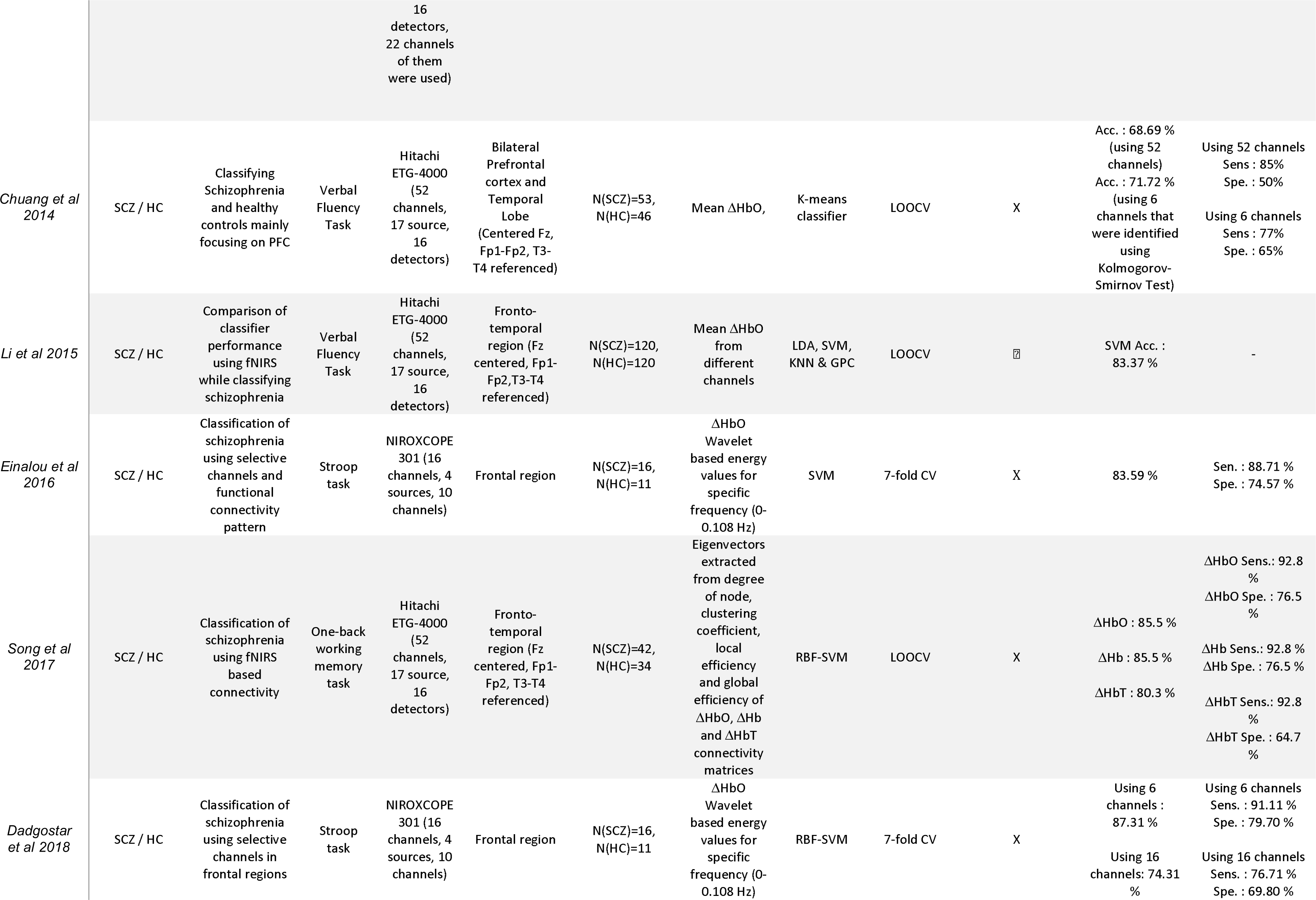
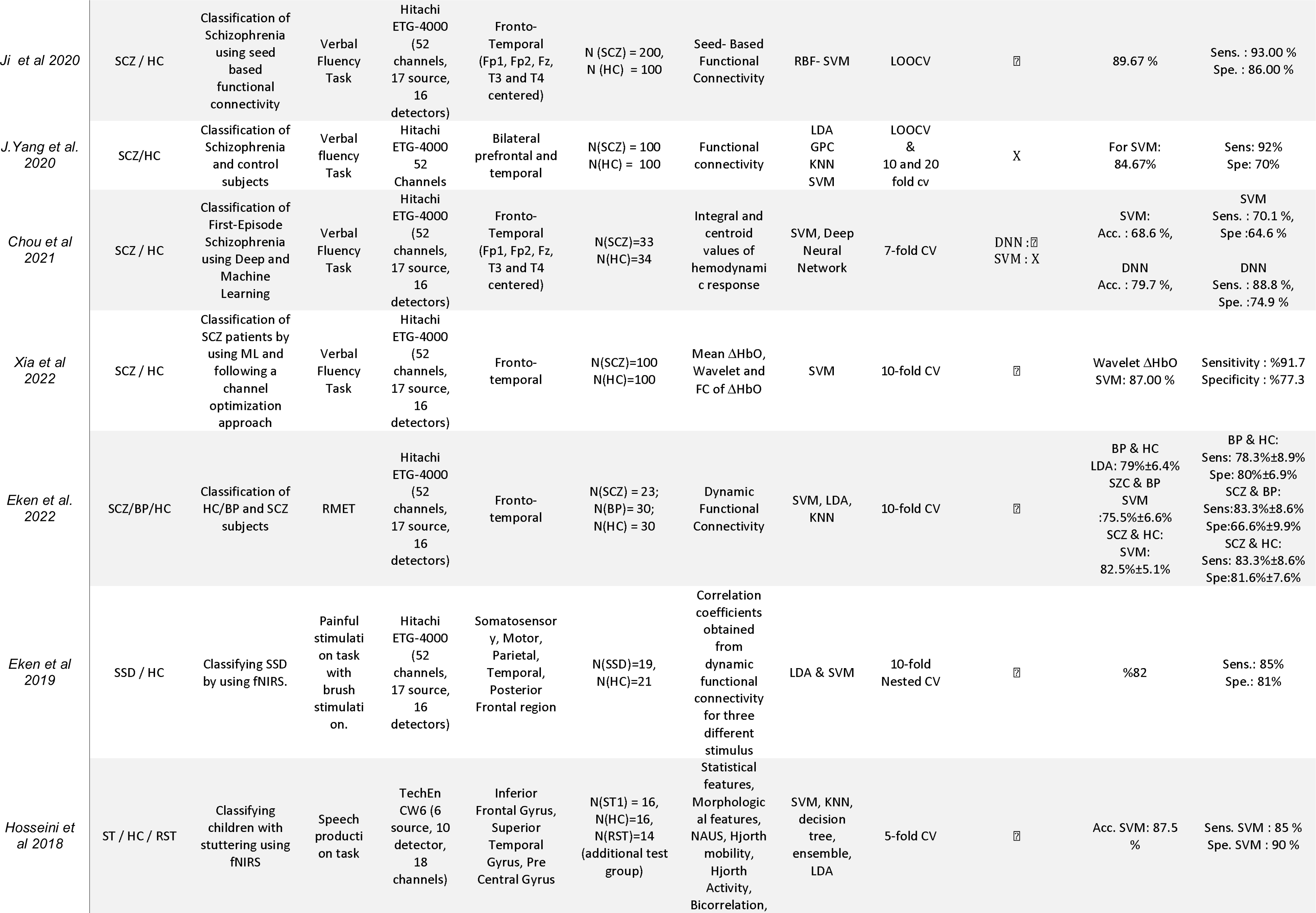
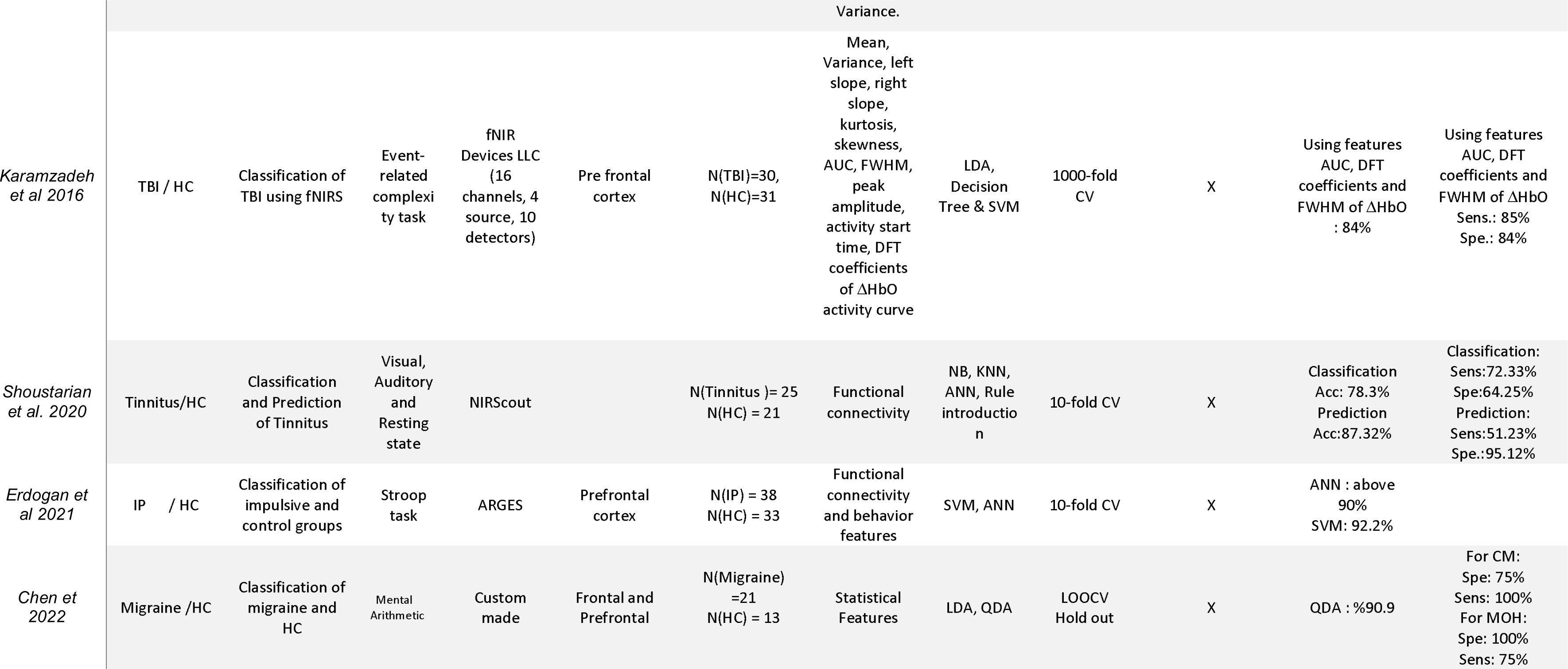
fNIRS studies that utilizes Machine Learning for clinical populations. Acc. : Accuracy, ADHD : Attention Deficit and Hyperactivity Disorder, ASD : Autism Spectrum Disorder, AUC : Area under curve, BP : Bipolar Disorder, CNN : Convolutional Neural Network, CV: Cross Validation, DFT: Discrete Fourier Transform, DLPFC : Dorsolateral Pre Frontal Cortex, EEG: Electroencephalography, FM: Fibromyalgia, FWHM : Full Width Half Maximum, GPC : Gaussian Process Classifier, HC: Healsl,thHyDRco:nHtermo odynamic response, IFC: Inferior Frontal Cortex, IP : Impulsive disorder, KNN : K-nearest neighborhood, L : Left, LA : Left Arm, LDA: Linear Discriminant Analysis, LF : Left Foot, LH : Left Hand, LOOCV : Leave-one-out cross validation, LR : Linear Regression, LSTM: Long-short term memory, Max. : Maximum, MCI : Mild Cognitive Impairment, MDD : Majo ri sDoerpdreers,sMi vFeGD: Middle Frontal Gyrus, MFC : Medial Frontal Cortex, MI : Primary Motor Cortex, Min. : Minimum, MLP : Multi-Layer Perceptron, MPH : Methylphenidate, MVPA : Multi-Voxel Pattern Analysis, NA: Not available, NA Normalized Area Under Signal, NPV : Negative Predictive Value, PFC: Pre-frontal Cortex, PPV : Positive PredicPt irveeCVeanlturea,l GP ryer uCsG,:R: Right, RA : Right Arm, RF : Random Forest, RBF : Radial Basis Function, RiF: Right Foot, RH : Right Hand, RST : Recovered from Stuttering, QDA : Qua nr itmit ai ntai vnet DAinsac lysis, SCZ : Schizophrenia, Sens. : Sensitivity, SI: Somatosensory Cortex, SMA : Supplementary Motor Area, Spe. : Specificity, SSD : Somatic Symptom Disorder, SST: Stop Signal Task, ST : Stuttering group, ST1 & 2: Stuttering group 1 & 2, SVM : Support Vector Machine, TBI : Traumatic Brain Injury, ΔHb : Deoxy-hemoglobin, ΔHbO : Oxy-hemoglobin.

### 2.3. Statistical Analysis

All statistical analyses and graphical representations were performed by using R (v4.1.2; R Core Team 2021). We performed Shapiro-Wilk test to control whether the data is normally distributed or not and applied correlation and correlation analysis between sample size and accuracy values.

## 3. Results

According to distribution of number of studies, for the last 13 years, using ML in fNIRS based clinical studies has an increasing trend. On the other hand, vast majority of these fNIRS based ML studies focused on SCZ (n=12), ADHD(n=7), ASD (n=6), MDD (n=5), MCI (n=4) and AD (n=3) populations. We also included studies and labeled as “other” from many different clinical populations such as Amyotrophic Lateral Sclerosis (ALS), Bipolar disorder (BP), Fibromyalgia (FM), Parkinson’s Disease (PD) Somatic Symptom Disorder (SSD), Stuttering, Traumatic Brain Injury (TBI) and Migraine. From 2010 to 2018, only four populations (SCZ, ADHD, TBI and stuttering) were studied. However, after 2019, more populations were also studied. Number of the studies per population for every year is shown in Figure 3.

**Figure 3.**
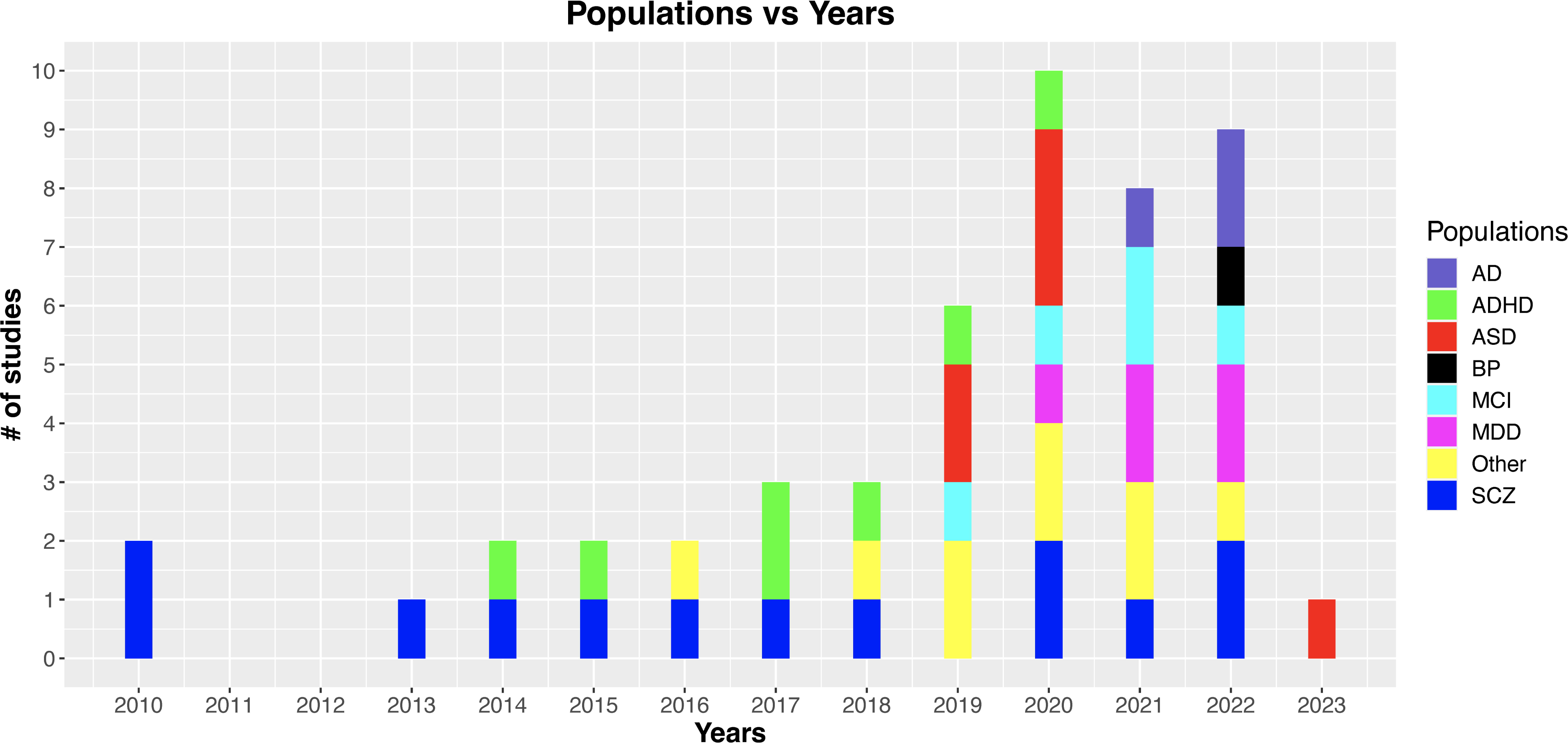
Number of fNIRS-based machine learning studies that includes clinical populations since 2010.

### 3.1. Attention Deficit and Hyperactivity Disorder (ADHD)

Among seven ADHD based classification studies, five of them focused on ADHD vs HC classification. Except for the studies that focused on only frontal region such as Güven and her colleagues (Güven et al., 2020) and Yasumura and her colleagues (Yasumura et al., 2017), all these studies focused on frontal and temporal region for classification. SVM is the most popular algorithm for ADHD / HC classification (n=5), except for two studies all studies used mean ΔHbO as feature, vast majority of studies used cross-validation method as LOOCV (n=4).

Vast majority of these studies have generally low sample sizes (min-max: 17-50) except for Yasumura and colleagues (Yasumura et al., 2017). This study is a multi-center study performed to validate the reliability of a classifier. It includes the highest number of subjects (Training data; ADHD: 108, HC: 108. Validation data; ADHD: 62, HC: 37) among all ADHD classification studies using fNIRS. fNIRS data that was acquired from PFC via a reverse Stroop task from different centers were used as input data with behavioral and physiological features. 86.25 % accuracy was found by using Radial Basis Function (RBF)-SVM and reverse stoop task-induced PFC activation was suggested as a critical biomarker for ADHD diagnosis. Accuracy values for other studies varies between 77.20 % - 86.00 % which could not exceed Yasumura and colleagues’ study despite their low sample sizes (Crippa et al., 2017; Gu et al., 2018; Güven et al., 2020; Ishii-Takahashi et al., 2015). On the other hand, in these studies, mean ΔHbO is the most popular feature for the classification in ADHD and also provides 86.25 % (Yasumura et al., 2017), 86.00 % (Gu et al., 2018) and 81.00 % (Ishii-Takahashi et al., 2015) accuracies which are the highest accuracies across all ADHD / HC classification studies. It can be interpreted that fronto-temporal region might provide critical biomarkers to distinguish ADHD and HC groups. However, more studies that follows similar procedures from experimental design to machine learning steps are needed.

In addition to this, two fNIRS studies focused on ADHD / ASD classification. One of those studies focused on hemodynamic biomarkers in the occipital region induced by a face-familiarity task, however, their sample size is relatively quite small (N=17, ADHD=9, ASD=8) compared to other ADHD classification studies and they found 84 % accuracy by using SVM(Ichikawa et al., 2014). The other study focused on the question that hemodynamic response after MPH medication and found 82 % accuracy after pooling results of six different classifiers (Simple, AND, OR, LDA, quadratic discriminant analysis, SVM) (Sutoko et al., 2019). Due to two different concepts of experiments and classification approaches, it is difficult to perform a comparison between the studies.

### 3.2. Alzheimer’s Disease (AD)

Among all AD (n=3) classification studies, Ho and colleagues’ study is the one the highest number of participants and they proposed a deep learning framework for sub-population classification of AD (T. K. K. Ho et al., 2022). 140 subjects including 53 HC, 28 asymptomatic AD, 50 prodromal AD and 9 AD dementia attended an fNIRS session focusing on prefrontal cortex. Highest accuracy was found as 90% ± 1.2%. Kim and colleagues also conducted a study to predict AD stages (J. Kim et al., 2022). 168 subjects (70 HC, 42 MCI, 21 Mild AD, and 35 moderate AD) were recruited and RF was used as classifier. 94.4 % accuracy was found to classify AD. Another study that tried to classify AD, MCI and HC subjects was conducted by Kim and colleagues (E. Kim et al., 2021). In this study, 60 participants (18 AD, 11 MCI and 31 HC) were recruited and PFC based FC of ΔHbO values were used as input of artificial neural network (ANN) classifier to classify disease state highest accuracy was found as 93.7%.

It is difficult to perform a direct comparison between studies due to the variability of sample size, different feature types and different classifiers. More studies are needed to make proper interpretation.

### 3.3. Autism Spectrum Disorder (ASD)

All reported ASD classification studies were done by using a similar dataset except for the study Dahan and colleagues performed (Dahan et al., 2020). 26 ASD patients were attended to the study to classify Autism Spectrum Quotient (AQ) patients according to their severity. The highest accuracy that was reached in this study was reported as 96.3% when RF was used as a classifier.

Rest of the studies were carried out by using the same dataset. In this dataset, 47 children (Typical developing (TD)=22, ASD = 25) were recruited and an 8 min of resting-state measurement from bilateral temporal regions was performed. In the first study (Xu et al., 2019), a convolutional neural network (CNN) with a gate-recurrent unit (GRU) was trained and tested via hold-out cross-validation and 92.2 % accuracy with 85 % sensitivity and 99.4 % specificity was found. Second study was performed by Cheng and colleagues (Cheng et al., 2019). In addition to the features used in the previous study, a specific frequency of interest for both ΔHbO (0.02 Hz) and ΔHb (0.0267 & 0.0333 Hz) in TC was also added as a feature and used as an input for an SVM classifier. With this new feature set, 92.7 % accuracy was found. The major difference between the two groups was reported as in the frequency band of 0.02-0.03 Hz. However, only a 0.5 % increase in accuracy was observed.

Sample entropy as a feature was also tested on the same dataset (Xu, Hua, et al., 2020). Using k-means classification, 97.6 % accuracy was found. After performing machine learning studies, two deep learning studies on similar data were recently reported (Xu et al., 2019; Xu, Liu, et al., 2020). In the other study (Xu, Liu, et al., 2020), CNN and long-short term memory (LSTM) were trained and tested via hold-out cross-validation and 95.7 % accuracy was reported. Another study that tries the diagnosis of ASD patients was conducted by Li and colleagues (C. Li et al., 2023). This study proposes a CNN-based algorithm by using resting-state fNIRS signals of 25 ASD children and 22 HC children. 12 channels located on frontal and temporal regions recorded NIRS signals by using FOIRE 3000 continuous NIRS system. Maximum accuracy that reported in this study is 94%.

Compared to deep learning approaches, a clustering based algorithm, k-means outperformed previously reported machine learning and deep learning results. This performance might also be due to the sample entropy which seems to be a potential biomarker to distinguish ASD and HC.

### 3.4. Mild Cognitive Impairment (MCI)

Among the four studies, three of them were published by the data using the same population (24 participants, MCI:15, HC :9). First study on MCI classification was performed by Yang and colleagues (Yang et al., 2019). 24 participants (15 MCI: 9 HC) were recruited for this study and statistical features of ΔHbO and ΔHb, activation t-maps and channel by channel correlation-maps were extracted. %90.62 accuracy were found by using convolutional neural network (CNN) and t-maps. Same group also performed another DL study that used and in addition to statistical features they also used ΔHbO spatio-temporal maps (D. Yang & Hong, 2020). Highest accuracy that was reached in this study was 98.61%. Last study by using the same population focused on transfer learning based classification of MCI and by using connectivity maps they found 97.01 % accuracy (Yang & Hong, 2021). This dataset has a low sample size to classify MCI and it is hard to interpret a general overview related to populations and applied methods.

In addition to this dataset, two studies include MCI populations in addition to AD population. First of these studies focused on FC of ΔHbO and tried to classify the MCI population (E. Kim et al., 2021).60 participants (18 AD, 11 MCI and 31 HC) were recruited and by using an artificial neural network (ANN) classifier they found 99.3 % accuracy for MCI classification. In the second study, 168 participants (70 HC, 42 MCI, 21 Mild AD, and 35 moderate AD) were recruited and 92.6% accuracy was found for MCI classification by using ΔHbO time series and random forest (RF) algorithm (J. Kim et al., 2022).

### 3.5. Major Depressive Disorder (MDD)

For MDD / HC classification, five studies have been reported. In the first study, 31 participants (14 HC and 17 MDD) were recruited and ten statistical features were extracted from ΔHbO of DLPFC and VLPFC and five of those features (ΔHbO variance from left DLPFC, mean ΔHbO from left VLPFC, FWHM of ΔHbO from medial PFC, mean ΔHbO from right VLPFC and Kurtosis of ΔHbO from right DLPFC) gave the highest accuracy for both XG Boost classifiers as 92.6 % (Zhu et al., 2020).

Similar statistical features are also used by Chao and colleagues (Chao et al., 2021) and they recruited 32 participants (16 MDD and 16 HC). By using statistical-based features with four vector-based features such as Cerebral Blood Volume (ΔCBV), Cerebral Oxygen Change (ΔCOE), angle K (ΔCOE/ΔCBV) and cascade forward neural network (CFNN), highest accuracy was achieved by using RNN and was found 99.86% by using only vector-based features. Also, this study claimed that AUC and angle K of fNIRS signals recorded from the prefrontal cortex (PFC) are specific neurological biomarkers for detecting MDD. Wang and colleagues recruited 96 subjects for MDD / HC classification (Wang et al., 2021) however, there is a great imbalance between classes (79 MDD and 17 HC subjects). Highest accuracy of 90% was achieved by using AlexNet model and correlation maps as input.

Highest number of participants were attended to the studies Li and colleagues (n=363, MDD=177, HC = 186) (Z. Li et al., 2022) and Ho and colleagues (n=133, MDD = 65, HC=68) (C. S. Ho et al., 2022). In both studies, verbal fluency task (VFT), which is a popular task in MDD research to reveal potential differences between MDD and HC groups (Henry & Crawford, 2005) were used. In both studies, SVM classifier were used and extracted features were integral and centroid values for Li and colleagues and FC of ΔHbO and ΔHb for Ho and colleagues. When compared the results of both studies, Li and colleagues found higher accuracy (75.6 %) than Ho and colleagues (73%).

On the other hand, when we analyzed the sample size and accuracy relationship for only MDD studies, there is a negative non-significant correlation is observed (r=-0.8, p=0.1). Due to the less number of studies, further studies are needed to clarify whether there is a significant trend between sample size and accuracy.

### 3.6. Schizophrenia (SCZ)

SCZ is the most studied population using fNIRS and ML approaches. In addition to conventional experimental studies since the first study published in 1994 (Okada et al., 1994), eleven machine learning studies have been performed by utilizing fNIRS since 2010. The vast majority of those studies focused on the prefrontal cortex (PFC) based on differences between two populations, most popular features was mean ΔHbO (n=5) and FC of ΔHbO (n=4) and most popular ML algorithm is SVM (n=8). There is not significant correlation between sample sizes and accuracy values for SCZ studies (r=0.11, p=0.74). Among 11 studies only 5 of them were able to recruit more than 100 participants (Azechi et al., 2010; Ji et al., 2020; Z. Li et al., 2015; Xia et al., 2022; J. Yang et al., 2020).

Among these four studies, the first study was performed by recruiting 120 participants (SCZ =60, HC =60) and 60 of them (30 HC, 30 SCZ) were used for training and testing a LDA classifier and the remaining participants (30 HC, 30 SCZ) were used for validation the LDA classifier (Azechi et al., 2010). Classification results by using only frontal mean ΔHbO showed a 78.3 % accuracy for the first group and for the second testing group, 65 % accuracy was observed. Li and colleagues recruited 240 participants (SCZ=120, HC=120) (Z. Li et al., 2015) and four different classifiers (LDA, SVM, KNN, GPC) were trained using the frontal mean ΔHbO. The highest accuracy was found by using Radial Basis Function (RBF) SVM (83.37 %). Ji and colleagues were able to recruit 300 (SCZ=200, HC=100) participants in their study (Ji et al., 2020) and utilized FC of ΔHbO for classification. They found 89.67 % accuracy in their study. Also, Yang and colleagues recruited 200 participants (SCZ=100, HC=100) and utilized FC strength of ΔHbO for classification like previous study (J. Yang et al., 2020) and they found 84.67 % accuracy. Xia and colleagues recruited 200 participants (SCZ=100, HC=100) and by using wavelet based features of ΔHbO and SVM, they found 87.00 % accuracy (Xia et al., 2022). Among these studies Ji and colleagues were able to find highest accuracy despite having a higher sample size. However, in general SVM based studies has higher accuracy compared to other classifiers (K-means, LDA, DL and other classifiers) (t(5)=4.838, p=0.010) despite not having statistically significant difference between their sample sizes (t(5)=1.693, p=0.131). In addition to efficiency of SVM, studies utilizing FC of ΔHbO provided greater accuracy than studies utilizing mean ΔHbO. Therefore, SVM and FC of ΔHbO might be an effective combination to accurately classify SCZ.

On the other hand, Hahn and colleagues recruited 80 participants (SCZ =40, HC=40), used whole ΔHbO response from fronto-temporal region and performed a classification study utilizing a probabilistic method (Hahn et al., 2013) and 76% accuracy was found. Chuang et al. also focused on PFC-based biomarkers in SCZ and tried to classify them using a k-means approach (Chuang et al., 2014). 99 participants (SCZ =53, HC=46) were recruited and mean ΔHbO was used as feature and highest accuracy was found as 71.72 % by using 6 channels located on left IFG (5 of them) and right IFG (one of them). PFC oriented specific channel selection approach was also used by Einalou and colleagues (Einalou et al., 2016). 27 participants (SCZ:16, HC :11) were recruited and by using wavelet transform, 0.003-0.11 Hz frequencies were found critical for classification and genetic algorithm was used to select channels in PFC. Using SVM, they found 83.59 % accuracy. Another wavelet based SCZ classification study was performed by Dadgostar and colleagues (Dadgostar et al., 2018). 27 participants (HC=11, SCZ =16) were recruited and frontal ΔHbO wavelet-based energy values for 0-0.108 Hz were extracted using WBD for 16 channels and channel selection was performed by using a genetic algorithm and this input was given an RBF-SVM classifier. 87.31% accuracy was reported by using only 6 channels. In addition to wavelet based features, Chou and colleagues utilized integral and centroid values of HbO response for classification (Chou et al., 2021). From 67 participants (33 first episode SCZ and 34 HC) integral and centroid values of oxyhemoglobin changes were computed from fNIRS signals during a VFT task. SVM and DNN were used as classifiers. DNN reached better accuracy than SVM, with 79.9% while SVM accuracy was 68.8%.

fNIRS-based functional connectivity was also considered as a biomarker in SCZ discrimination (Song et al., 2017). 76 participants (SCZ =42, HC=34) were recruited and activity from the frontotemporal region was recorded. After creating connectivity matrices for ΔHbO, ΔHb and ΔHbT, eigenvectors extracted from the degree of node, clustering coefficient, local efficiency and global efficiency of three concentration changes were extracted as features and given as input to RBF – SVM classifier. Higher accuracies were reported by using ΔHbO and ΔHb (85.5 %) compared to ΔHbT (80.3 %). In another connectivity based classification study, Eken and colleagues utilized dynamic functional connectivity of ΔHbO to classify SCZ (Eken et al., 2022). 83 participants (23 SCZ, 30 BP and 30 HC) attended to fNIRS recording session during reading the mind in the eyes (RMET) task. By using SVM, highest accuracy was found as 82.5 %.

### 3.7. Other Populations

Nine studies were included in this group focusing on populations from Amyotrophic Lateral Sclerosis (ALS), Bipolar disorder (BP), Traumatic Brain Injury (TBI), Tinnitus, Stuttering, Somatic Symptom Disorder (SSD), Migraine, Parkinson’s Disease (PD), Fibromyalgia (FM) and impulsivity. Sample size varies between 18-71 and found accuracy values were between 62.64 - 100 %. Among these studies, vast majority of studies utilized SVM (n=5) as classifier, K-fold (n=7) as cross-validation approach and used statistical features of ΔHbO (n=3) and FC of ΔHbO (n=3) and only two studies performed hyperparameter optimization for classification.

For ALS classification, Deligani and colleagues performed a classification by using peak value and AUC of ΔHbO and SVM as classifier.(Deligani et al., 2021). 18 participants (9 ALS, 9 HC) were recruited and 62.64% accuracy was found by using only fNIRS-based features. Eken and colleagues (Eken et al., 2022) also performed a classification to classify Bipolar disorder by recruiting 60 participants (30 BP and 30 HC) and FC of ΔHbO was used as feature. Highest accuracy was found by using SVM algorithm as 82.5 %. Karamzadeh and colleagues performed TBI classification by recruiting 61 participants (TBI =30, HC =31) (Karamzadeh et al., 2016). Statistical features of ΔHbO were extracted and, the highest accuracy was found as 84 % by using AUC, DFT coefficients and FWHM of ΔHbO activity and decision tree classifier. Shoustarian and colleagues published a Tinnitus classification study by recruiting 46 participants (Tinnitus =25, HC = 21) (Shoushtarian et al., 2020). FC of ΔHbO and ΔHb were used as features and highest accuracy was found as 78.3% by using NB classifier. Hosseini and colleagues performed a stuttering classification study by recruiting 32 children (stuttering :16, HC:16) (Hosseini et al., 2018). Statistical features were extracted from ΔHbO and highest accuracy was found by using SVM as 87.5 %.

Eken and colleagues performed the first classification study on SSD population (Eken et al., 2019). 40 participants (HC=21, SSD = 19) were recruited FC of ΔHbO was used as feature 82% accuracy was found by using SVM classifier. Chen and colleagues conducted a study to classify migraine levels (Chen et al., 2022). 34 participants (13 HC, 9 chronic migraine patients (CM), 12 medication-overuse headache patients (MOH)) were attended to this study. Time domain feature extraction methods were performed on HbO and HHb signals in addition to total hemoglobin (HbT) and oxygen exchange (COE). Quantitative Discriminant Analysis (QDA) was used for classification and 90.9% accuracy was found for migraine / HC classification.

PD classification study using fNIRS and EEG was conducted by Abtahi and colleagues (Abtahi et al., 2020). 18 participants (PD:9, HC:9) were recruited and by using only mean ΔHbO, 81.23 % accuracy were found by utilizing SVM classifier. Gokcay and colleagues performed a FM classification study using likelihood-based decision level fusion approach of several classifiers (Gokcay et al., 2019). 36 participants (19 FM and 17 HC) were recruited and SVM, K-nearest neighborhood (KNN), and Linear Discriminant Analysis (LDA) with different parameters were trained and tested. After fusing the decision, 100 % accuracy was found. Erdogan and colleagues proposed a computer-based decision support approach for impulsivity classification (Erdogan et al., 2021) 71 participants (38 impulsive adolescents and 33 HC) were attended to this study and connectivity-based features were extracted from fNIRS signals and 61.6 % accuracy was found by using SVM classifier.

### 3.8. Sample Size and Accuracy

Effect of sample size on accuracy was shown in Figure 4. Among these included studies only 8 of them has more than 100 samples. 14 of these studies has sample sizes between 50 and 100 and the rest of the studies has sample size lower than 50. To find the statistical relationship between sample size and accuracy, first we checked whether our sample size and accuracy values were normally distributed and found that while our sample size data was not normally distributed (W=0.685, p=5.904*10^-9^), accuracy values data was normally distributed (W=0.965, p=0.15). We performed Spearman’s rank correlation to understand the relationship between the sample size and accuracy and found that there is no significant correlation between them (r=-0.24, p=0.09). However, when we exclude the studies that have lower sample size than 20, we found a negative significant correlation between the sample size and accuracy (r=-0.38, p=0.009).

**Figure 4.**
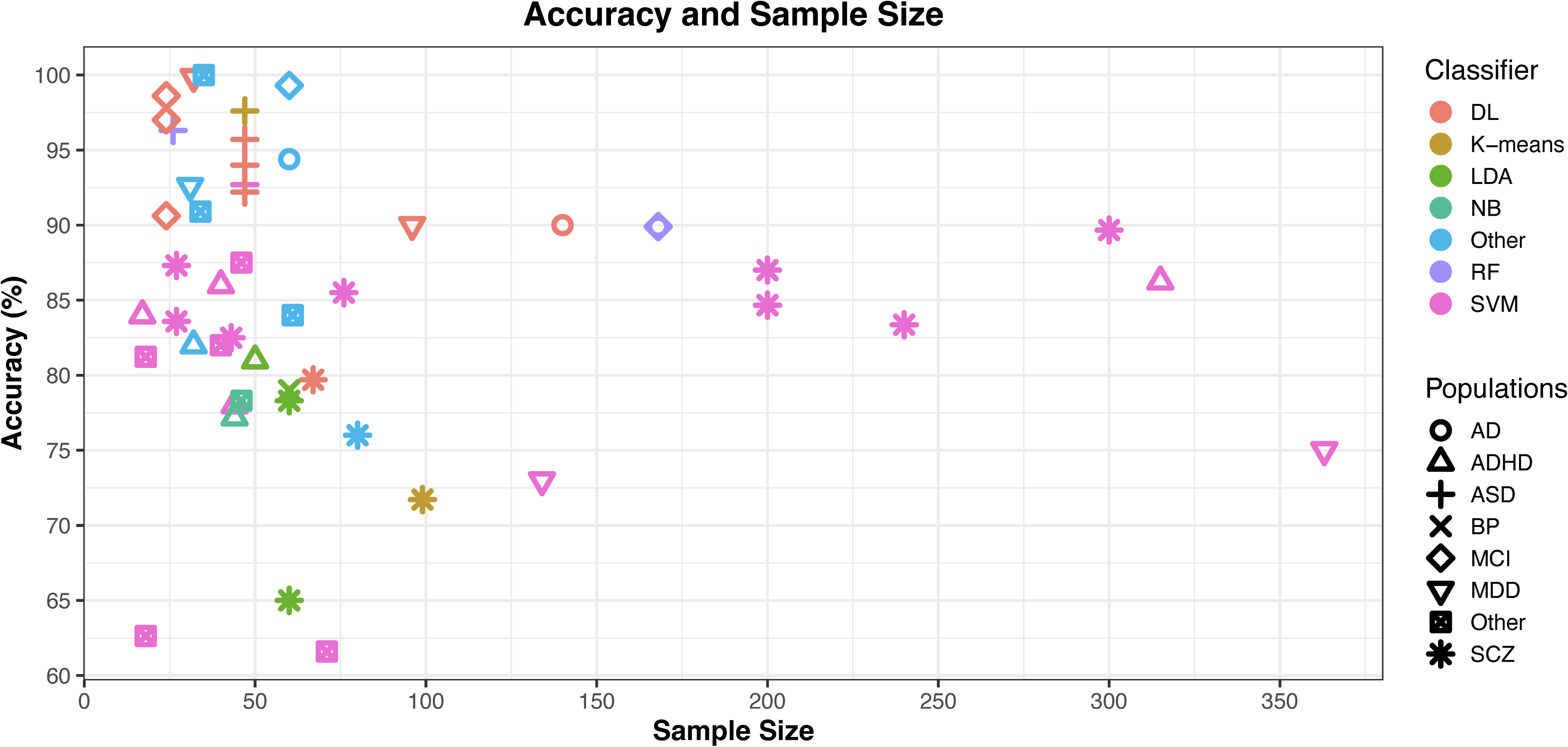
Accuracy values vs Sample size distribution with respect to classifiers and populations. DL: Deep Learning, LDA: Linear Discriminant Analysis, NB: Naïve Bayes, RF: Random Forest, SVM: Support Vector Machine, AD: Alzheimer’s Disease, ADHD: Attention Deficit and Hyperactivity Disorder, ASD: Autism Spectrum Disorder, BP : Bipolar Disorder, MCI: Mild Cognitive Impairment, MDD: Major Depressive Disorder, SCZ: Schiophrenia

When we perform the correlation analysis for the populations SCZ, ADHD, ASD, MDD and MCI separately, we found that there is no significant correlation between accuracy and sample size for ADHD (r=-0.018, p=0.97), ASD (r=-0.39, p=0.44), MCI (r=-0.22, p=0.72), SCZ (r=0.11, p=0.74) and MDD (r =-0.8, p=0.13).

### 3.9. Classifiers

Many different machine learning algorithms were used in fNIRS studies. Majority of fNIRS studies uses SVM (n=20), DL (n=10) methods and LDA (n=4) as classifiers. Distribution of classifiers and used populations are shown in Figure 5.a. SVM is an effective algorithm for low sample size and provides notable accuracy values even in high sample sizes and accuracy values were found between 61.60% - 92.70 % in studies published between Since 2014 to 2022. SVM classifier was used in study to classify populations ADHD(n=3), ADHD/ASD (n=1), ASD(n=1), ALS(n=1), MDD(n=2), PD(n=1), SCZ(n=7), SSD(n=1), ST(n=1) and impulsivity(n=1). In studies that uses SVM, sample size varies between 17 and 363.

**Figure 5.**
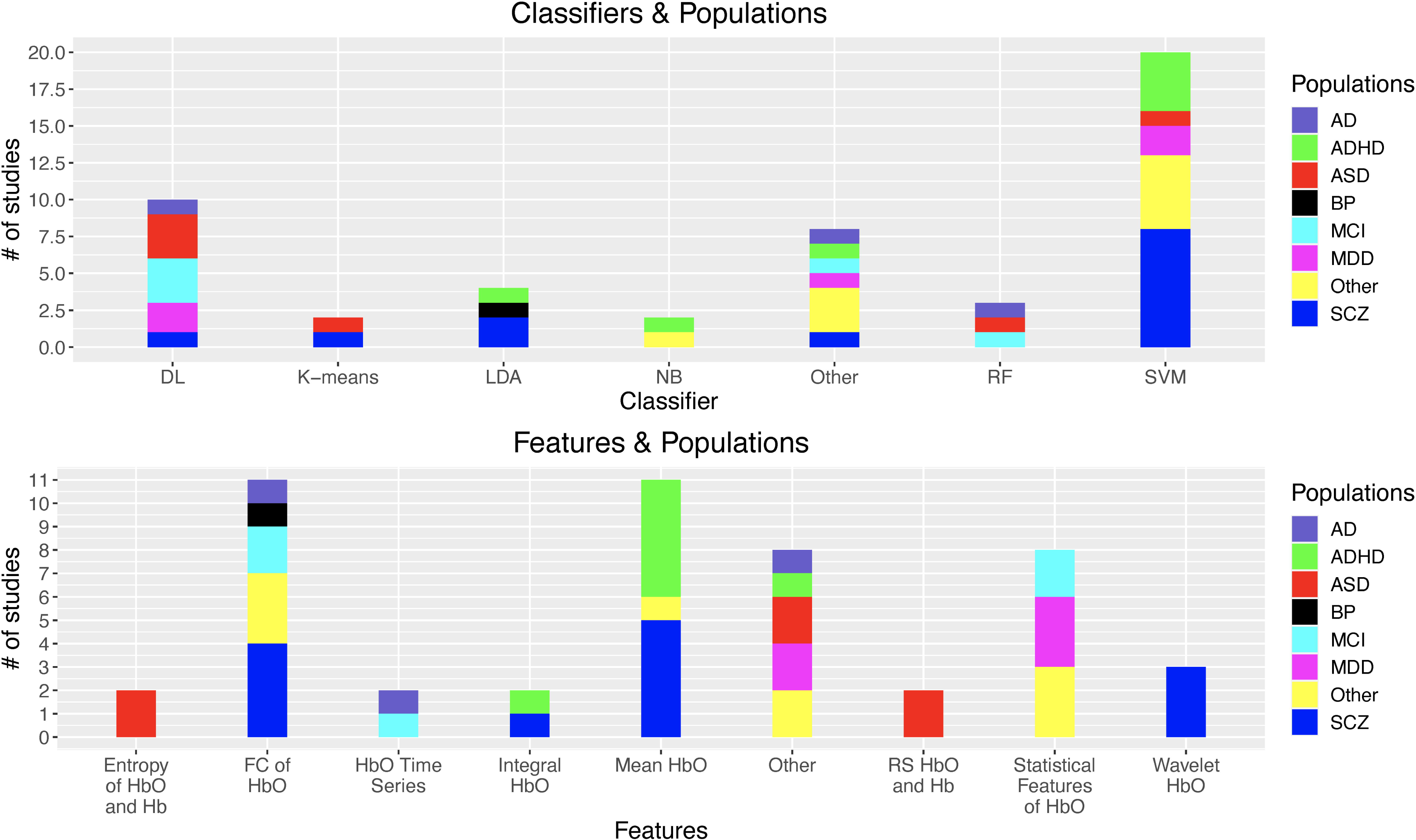
a) Distribution of number of studies with respect to classifiers and populations. b) Distribution of number of studies with respect to features and populations. DL: Deep Learning, LDA: Linear Discriminant Analysis, NB: Naïve Bayes, RF: Random Forest, SVM: Support Vector Machine, AD: Alzheimer’s Disease, ADHD: Attention Deficit and Hyperactivity Disorder, ASD: Autism Spectrum Disorder, BP : Bipolar Disorder, MCI: Mild Cognitive Impairment, MDD: Major Depressive Disorder, SCZ: Schiophrenia. HbO : Oxy-hemoglobin concentration change (HbO, Hb: Deoxyhemoglobin concentration change. RS: Resting State.

On the other hand, second greatest classifier group is DL based methods. DL based methods require big data due to tuning the weights of methods during training session. However, in recent years data augmentation methods (adding gaussian noise, spikes, trend) on time series were used to increase the number of training samples after separating the validation and test datasets (Iglesias et al., 2023). DL based classifiers were applied to populations AD(n=1), ASD (n=3), MDD(n=2), MCI(n=3), SCZ (n=1) and accuracy values vary between 79.9 % - 98.61 %.

### 3.10. Feature Engineering

In this review, feature types can be grouped under three different categories; time series based features such as mean ΔHbO and statistical features such as mean, std, kurtosis, skewness, slope and functional connectivity-based features. Most popular features in these studies were functional connectivity by using ΔHbO (n=11), mean ΔHbO (n=11) and statistical features such as std. dev, variance, skewness which are generally used in BCI studies (n=8). Distribution of features with respect to populations are shown in Figure 5.b. Connectivity-based features have also emerged as another alternative input for ML algorithms. Due to its nature, resting-state-based classification studies using fNIRS utilize these features (Cheng et al., 2019; J. Li et al., 2016; Xu et al., 2019; Xu, Liu, et al., 2020). In addition to this, some task-based studies also use connectivity-based features (Eken et al., 2019; Gokcay et al., 2019; Song et al., 2017; Yang et al., 2019).

### 3.11. Optimizing Hyperparameters

Hyperparameter optimization were performed only for 16 studies. In Figure 6.a. number of studies that applied parameter optimization with respect to classifiers are shown. To improve the performance of classifiers, optimizing hyperparameters using different approaches is an option. Vast majority of parameter-optimized classification studies used Grid-search parameter optimization (Z. Li et al., 2015; Yang et al., 2019; Yasumura et al., 2017) and Bayesian optimization (Eken et al., 2019; Hosseini et al., 2018). The grid-search algorithm creates all combinations of parameters and trains the classifier by using these parameters. After training all, it gives the optimum parameter set that provides the lowest validation error. Grid-search is computationally expensive both for time and space. Also, as the number of parameters increases, computational complexity becomes high. On the other hand, Bayesian optimization is a sequential iterative optimization process that aims to find the global optimum set of parameters using minimum iterations. Compared to grid search, it uses less training time but, considers fewer options. For deep learning studies, Adam (adaptive moment estimation) optimizer is the most popular method for parameter optimization and is generally preferred in several fNIRS-based deep learning studies (Xu, Liu, et al., 2020; Yang et al., 2019).

**Figure 6.**
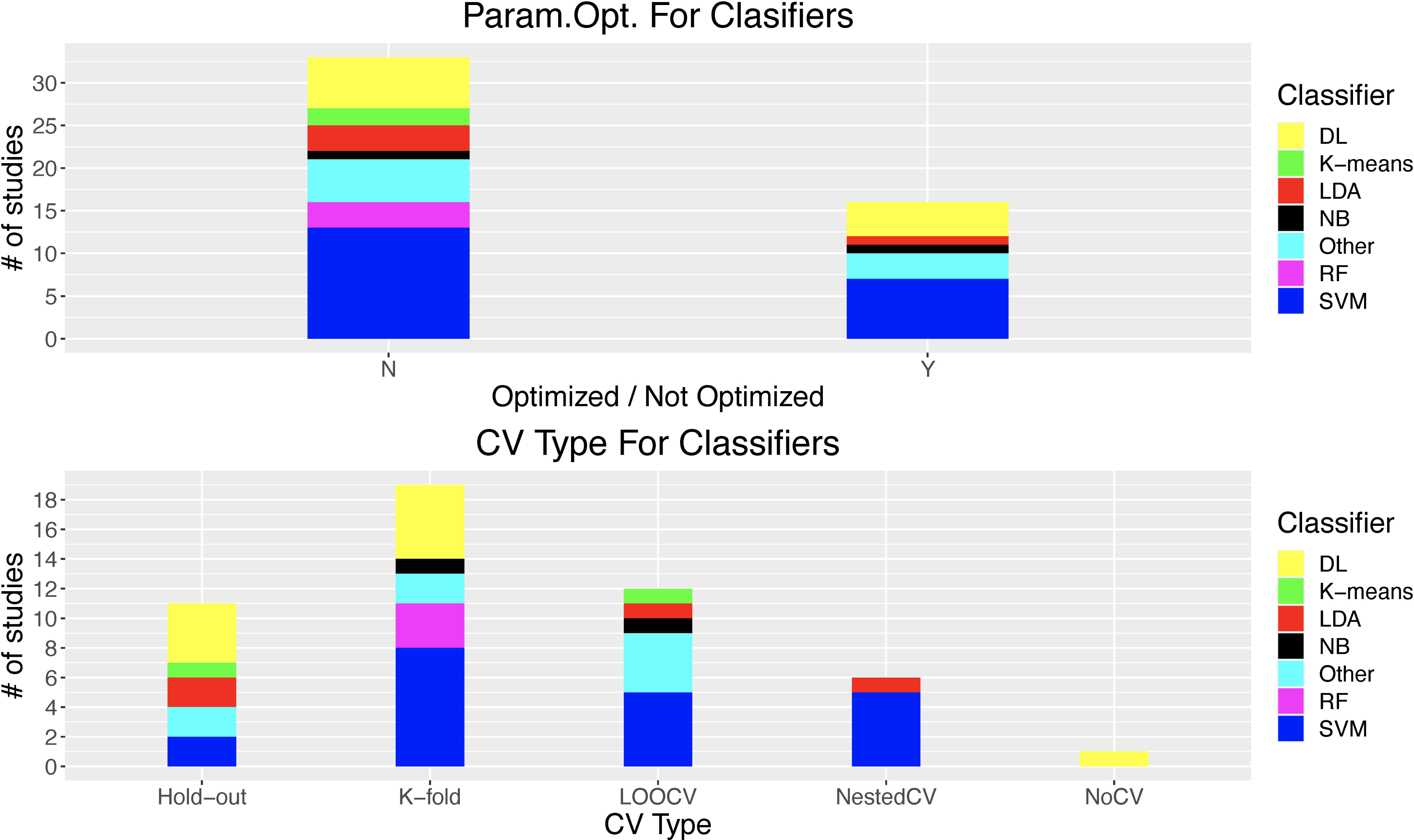
a) Hyperparameter optimization of classifiers and b) Applied cross-validation types to classifiers. . DL: Deep Learning, LDA: Linear Discriminant Analysis, NB: Naïve Bayes, RF: Random Forest, SVM: Support Vector Machine, Y: Optimized, N: Not optimized. LOOCV: Leave-one-subject-out cross-validation, Nested CV: Nested Cross-Validation

### 3.12. Cross-Validation (CV) Techniques

Most applied cross-validation types are k-fold cross-validation (n=18), leave-one-out cross validation (n=12) and hold-out cross validation (n=11) and Nested Cross-validation (n=6) In Figure 6.b. number of studies that applied cross-validation with respect to classifiers and cross-validation type are shown. We found that K-fold CV is the most popular CV method. In this method, observations are divided into K number of training and test folds that both training and test folds were stratified. For every fold, a classifier is trained by using training fold and tested by using test fold. This is done by K times. After having a classification score from every classifier, all these scores were averaged. It is ideal for moderate-sized (e.g.(N ≈ 50 - 100)) datasets. However, for larger datasets, it causes computational complexity. In this review, we saw that studies that have various number of samples used K-fold cross validation (min-max : 17 – 315).

In LOOCV, only a single observation from data is used for the test and the rest is used for training. This operation was done for every observation. Therefore, you have n test scores and then the average score is estimated. It provides less bias since all data is used for testing. However, for the same reason, variation is high in scores. Also, for larger samples (e.g. > 100-1000) computational cost is high. For 12 studies that used LOOCV, sample sizes were between 40-300 and the accuracy values were between 71.72 – 99.30 %.

For hold-out CV, data is separated as training and test set. Percentages vary around for training 60-90 % and test 10-40 %. Training and testing are done only once. This is ideal for a large dataset that requires more computational power and time. However, results are highly biased due to less generalization because training and testing samples might not represent the whole data. In this review, 11 studies that used hold-out CV have sample sizes lower than 100. These studies have generally higher accuracies (min 65 % - max 97.6 %). Also, among these 11 studies, 4 of them used deep learning which requires more data compared to conventional ML methods to adjust its weights depending on its size.

For some studies, nested CV is also used (Crippa et al., 2017; Eken et al., 2019). Nested CV consists of two nested loops. The outer loop is always for generalization of ML models and the inner loop is either for hyperparameter optimization or rarely feature selection (Parvandeh et al., 2020). It is used for having an unbiased estimate of classification scores. To optimize classification results with unbiased results, nested CV is a highly reliable approach. We have 6 studies that used Nested CV which have sample sizes between 40 - 363 and accuracy values were between 73-82.5 %.

## 4. Discussion

In this review, we analyzed the studies focusing of diagnostic ML applications by using fNIRS data. Compared to fMRI and EEG, few number of studies were published on diagnostic ML applications by using fNIRS. While several systematic reviews for diagnostic classification of SCZ (de Filippis et al., 2019; Shim et al., 2016) or ASD (Santana et al., 2022) were published by using fMRI or EEG, to our best knowledge this is the first review that focuses on diagnostic classification of disorders by using fNIRS and ML. Due to having similar features, fNIRS also shares the similar problems with other neuroimaging modalities.

### 4.1. Sample Size

Sample size is a chronic problem not only in conventional neuroimaging studies but also for ML applications. Among reviewed studies, only 8 of 45 studies have sample size greater than 100. In a recent review that covers 200 papers on diagnostic ML applications by using fMRI revealed that majority of these studies have sample size less than 150 (Arbabshirani et al., 2017). In a recent review it was reported that 300 neuroimaging studies published between 2017 and 2018, have sample size around 23-24 (Szucs & Ioannidis, 2020). Low sample size in neuroimaging studies led to several problems in replicability (Turner et al., 2018), cause high variance (Mumford, 2012) and low sample size with circular analysis cause higher classification accuracies which is possibly a misleading signature for diseases such as ADHD (Pulini et al., 2019). Also, applied cross-validation will cause a large error bias when the sample size is low (Varoquaux, 2018). Previous studies reported that low sample size-based classification studies reach higher accuracy when higher sample sizes lead lower accuracies (Schnack & Kahn, 2016).

To overcome sample size problem, first we think that fNIRS databases needs to be created. OpenfNIRS (https://openfnirs.org/data/), NITRC (https://www.nitrc.org/projects/fnirsdata/) were the only initiatives that allows sharing fNIRS data among researchers until now. However, few number of datasets are available in these databases and vast majority of these datasets include motion artifacts to test motion artifact correction methods. More specific population based databases needs to be created. Compared to fNIRS, there are several fMRI and MRI databases such as Alzheimer’s Disease Neuroimaging Initiative (ADNI) (Jack et al., 2008), openfMRI (Poldrack et al., 2013; Poldrack & Gorgolewski, 2017). Databases allow the researchers to reach big datasets and train and test their models. Like databases, more multi-center data collection should also be performed to generalize the performance of ML for diagnostic purposes. Until now, only one ML based multi-center studies were reported for ADHD (Yasumura et al., 2017).

Another problem related to sample size is data standardization. It is a great necessity to standardize some critical procedures such as anatomical positioning on common templates such as MNI (Tsuzuki et al., 2007). At this point, either utilizing MRI data of subjects or using 3D digitizers can be considered valid options to perform an accurate channel localization (Tsuzuki & Dan, 2014). Also, to assess regional biomarkers for every individual, cortical ROIs should be precisely defined and corresponding coordinates of this ROI should be reported. Some toolboxes provide anatomical information of channels by using MRI or 3D optode coordinate data such as AtlasViewer (Aasted et al., 2015), NIRS-SPM (Ye et al., 2009), NAP(Fekete et al., 2011a, 2011b) and fOLD (Zimeo Morais et al., 2018). This also will gain insight into further studies particularly comparing the results. For big datasets, datasets with a standard near-infrared data format .snirf (https://github.com/fNIRS/snirf) that includes spatial information are necessary. Many systems (NIRx, Kernel, Cortivision, Gowerlabs, Artinis) allows the researchers to export data in .snirf format. Therefore, not only the ML based classification or prediction studies related to specific disorders but also meta-analyses might be realized.

In this review, we found that there is a negative correlation between sample size and accuracy. A similar result was previously reported another review which focuses on deep learning studies on psychiatric populations using neuroimaging approaches (Quaak et al., 2021). Sample size has a great effect on classifier performance and higher sample sizes may include disease inhomogeneity therefore they can represent the whole population (Arbabshirani et al., 2017). After having enormous amount of high-quality data with accurate and precise spatial information, it will be possible to develop more accurate ML models for diagnostic purposes. a very common problem in low sample size and high dimension datasets is; they tend to cause overfitting if a proper feature selection is not done (Pereira et al., 2009).

### 4.2. Selected Features

For ML studies, the vast majority of the studies reported performance results by utilizing ΔHbO. However, notable number studies also considers about ΔHb as a critical feature source (Cheng et al., 2019; Chiarelli et al., 2021; Crippa et al., 2017; J. Li et al., 2016; Parent et al., 2019; Song et al., 2017; Sutoko et al., 2019; Xu et al., 2019; Xu, Hua, et al., 2020; Xu, Liu, et al., 2020; Xu et al., 2021; Yang et al., 2019; D. Yang et al., 2020). While selecting features for model training, ΔHbO based features are preferred for fNIRS analysis due to its high SNR compared to ΔHb (Homae et al., 2010; Montero-Hernandez et al., 2018; Niu et al., 2011; Zhang et al., 2010). It is also preferred in BCI studies (Naseer & Hong, 2015). However, some surprising results can be encountered such as finding higher accuracy by using ΔHb than using ΔHbO (Crippa et al., 2017; Xu et al., 2019). This is a controversial issue. Although there are some exceptional cases (Strangman et al., 2002), common agreement is that decrease in ΔHb is highly correlated with blood-oxygenation-level-dependent (BOLD) signal (Mehnert et al., 2013; Steinbrink et al., 2006). ΔHbO has a generally larger amplitude than ΔHb (Franceschini et al., 2000; Hirth et al., 1996; Shtoyerman et al., 2000). Due to this, ΔHb is easily affected by optical measurement errors (Strangman et al., 2002) which possibly might create false positive results in either conventional statistical analysis or machine learning results. However, on the other hand, recent evidence showed that ΔHb is less sensitive to extra-cerebral physiological noise interference and is found positively correlated to BOLD signal (Gervain et al., 2011; Mehnert et al., 2013; Steinbrink et al., 2006). There is no general consensus about the answer of the question which chromophore (ΔHbO or ΔHb) represents true hemodynamic behavior than the other. Due to this, we suggest that both signals should be considered as potential feature sources. In some cases, depending on the measure, ΔHb might provide better classification accuracies compared to ΔHbO (Crippa et al., 2017; Eken, 2021).

We also found that mean ΔHbO, FC of ΔHbO and statistical features were the most utilized features extracted from ΔHbO time series. A recent study comparing the performances of different features for MCI classification, found that, mean ΔHbO yielded higher accuracy than FC of ΔHbO (Xia et al., 2022). This is the only study that we were able to find such a comparison for a similar clinical group. However, this may change depending on the population, used algorithm, cross-validation type and many other factors. To interpret more generalizable results, more feature type comparison oriented studies are needed on specific clinical population datasets.

### 4.3. Cross-Validation and Hyperparameter Optimization

Cross-validation (CV) is a highly critical procedure for model generalization. After training the model, it should be tested on a separate different dataset or preferably validated and tested by using different datasets. However, due to data scarce which is often observed in neuroimaging studies, this generally might not be feasible. Only few studies applied an external dataset from a different cohort or site to test the model (Azechi et al., 2010; Hosseini et al., 2018; Yasumura et al., 2017). While determining the which CV type is used in studies, there are two aspects that needs to be considered bias/variance problem and model performance. In this review, three main CV technique are used. Leave-one-out cross validation (LOOCV), Hold-Out CV and K-fold CV. In LOOCV, only a single observation from data is used for test and the rest is used training. This operation was done for every observation. Therefore, you have n test scores and then average score is estimated. It provides less bias since all data is used for testing. However, for the same reason, variation is high in scores. Also, for larger samples (e.g. > 100-1000) computational cost is high. For hold-out CV, data is separated as training and test set. Percentages vary around for training 60-90 % and test 10-40 %. Training and testing are done only once. This is ideal for large dataset which requires more computational power and time. However, results are highly biased due to less generalization because training and testing sample might not represent the whole data.

Another popular CV method is Nested CV. It is generally preferred to perform either automatic feature selection or hyperparameter optimization (Arbabshirani et al., 2017). Among reviewed studies, studies that used nested CV (n=6) found accuracy values between 73-82.5 %. In these studies, vast majority of studies used SVM (Crippa et al., 2017; Eken et al., 2022; Eken et al., 2019; C. S. Ho et al., 2022; Z. Li et al., 2022). Vabalas and colleagues revealed that k-fold showed strongly biased performance with small sample sizes and nested CV produced robust and unbiased performance regardless of sample size (Vabalas et al., 2019). Nested CV is a computationally intense approach because it includes two nested loops and the pseudocode of nested CV is;

- *Divide the dataset into k folds,*
- *For each fold k_out=1….k: this is the outer loop for the generalization of classifier for to the selected hyperparameter*

○ *“Test_out” is the fold k_out, “Train_out” is the data except for other “Test_out” in fold k_out.*
○ *Divide the “Train_out” data into 10 folds*
○ *For each fold k_in2=1….k: this is the inner loop for the hyperparameter optimization.*

- *By using “Train_out” data, “Test _in2” is the fold k_in2,*
- *“Train_in2” is the data except for “Test_in2”.*
- *Divide the “Train_in2” into 5 folds*
- *Use “Train_in2” with each hyperparameter that was defined and evaluate it by using “Test_in2” and save the performance metrics.*
○ *Check the average score of each parameters over k-folds and choose the best one.*
- *Train the model with the best parameters by using “Train_out” and test it by using “Test_out”. Save the scores*.
- *Find the average scores by using all k folds*.

On the other hand, hyperparameter optimization approaches was utilized to improve model performances in only 16 studies. In some studies, without applying nested cross validation hyperparameter optimization was carried out by following k-fold cross validation (Yasumura et al., 2017). For DL studies, almost all of the studies utilized hyperparameter optimization. When hyperparameter optimization was not carried out, hyperparameters of classification algorithms (e.g. regularization parameter (C) of SVM, distance type of K-nearest neighbourhood) were randomly selected in other studies without justification and this bias might have affected performance of models.

To optimize hyperparameters for classifiers, grid-search, random-search and Bayesian search are the most popular optimization algorithms. In this review, among the all optimization algorithm vast majority of the studies uses grid-search optimization (Güven et al., 2020; Ji et al., 2020; E. Kim et al., 2021; Z. Li et al., 2015; Xia et al., 2022; Yasumura et al., 2017; Zhu et al., 2020) and Bayesian optimization (Eken et al., 2022; Eken et al., 2019; Hosseini et al., 2018). Among these algorithms, grid-search are computationally expensive due to the fact that as number of hyperparameters increases, number of trained models increases. However, it provides the best result among the all trained models depending on the given hyperparameter search space. For random-search, only a randomly selected part of given hyperparameters are searched. This approach is much faster than grid-search however, it does not guarantee the best result. Compared to grid-search and random-search, Bayesian search is an iterative method which selects its parameter set by considering the previous round score instead of randomly selecting a parameter set as random-search did or searching whole parameter set combinations as grid-search did. We suggest that if the aim is to obtain the best accuracy result regardless of its training time, grid-search is a better choice due to providing the best performance.

### 4.4. Limitations

There are several limitations in this review. First, compared to other neuroimaging modalities, few number of studies are reported. Several reviews were published related to diagnostic abilities of functional neuroimaging techniques such as fMRI (Arbabshirani et al., 2017; Bondi et al., 2023; Santana et al., 2022), EEG (Shim et al., 2016), PET (Duffy et al., 2019) and their interaction to machine learning approaches.

Studies generally reports multiple results, we extracted the best results among the results in a study. While reporting the studies, we basically focused on accuracy as the performance metric. While analyzing the studies, we generally focused on sample sizes, feature engineering and ML performance. However, there are also several critical factors that needs to be considered such as experimental design, focused ROI and data pre-processing pipelines of fNIRS signals. A recent study that compares different pre-processing approaches revealed that ignoring removal of task-evoked physiological noise led to different statistical results (Pfeifer et al., 2017). Also, a recent review showed that there is a high variability among pre- processing methods carried out in fNIRS studies (Pinti et al., 2018). These factors should also be considered in future reviews.

## 5. Conclusion

To our best knowledge, this study is the first review that focuses on diagnostic ML applications of fNIRS. fNIRS has been continuously gaining importance in neuroscience research due to its notable advantages compared to other modalities. On the other hand, its translation to clinics as a diagnostic tool is a highly critical research field. Nowadays, as we are experiencing AI age, its interaction to fNIRS is inevitable. While it is still in early stages, there are several promising results that were reported by utilizing this cooperation.

It is a widely known fact that fNIRS has several challenges such as data standardization, lack of data, and preprocessing problems. However, despite these pitfalls, there is a growing interest to understand the potential biomarkers to be used as discriminative parameters for different populations via fNIRS by utilizing ML approaches. In case of overcoming these problems mentioned above, ML diagnosis by utilizing fNIRS data for diagnostic purpose will have two benefits; 1) A critical decision support system for diagnosis without considering any subjective measure, 2) Suggesting potential biomarkers on cortical-regions for specific disorders that previously were not considered for diagnosis and compared to fMRI, these biomarkers might be more easier to reach.

## Data Availability

All data produced in the present study are available upon reasonable request to the authors.

## Acknowledgement

We would like to thank to Prof. Dr. Turgut Durduran from the Institute of Photonic Sciences (ICFO, Barcelona, Spain) for his valuable and constructive suggestions during the planning and development of this review.

## Data Availability Statement

No new data were created or analyzed in this study

